# The Maximal Expected Benefit of SARS-CoV-2 Interventions Among University Students: A Simulation Study Using Latent Class Analysis

**DOI:** 10.1101/2024.11.04.24316707

**Authors:** Callum R.K. Arnold, Nita Bharti, Cara Exten, Meg Small, Sreenidhi Srinivasan, Suresh V. Kuchipudi, Vivek Kapur, Matthew J. Ferrari

## Abstract

Non-pharmaceutical public health measures (PHMs) were central to pre-vaccination efforts to reduce Severe Acute Respiratory Syndrome Coronavirus 2 (SARS-CoV-2) exposure risk; heterogeneity in adherence placed bounds on their potential effectiveness, and correlation in their adoption makes assessing the impact attributable to an individual PHM difficult. During the Fall 2020 semester, we used a longitudinal cohort design in a university student population to conduct a behavioral survey of intention to adhere to PHMs, paired with an IgG serosurvey to quantify SARS-CoV-2 exposure at the end of the semester. Using Latent Class Analysis on behavioral survey responses, we identified three distinct groups among the 673 students with IgG samples: 256 (38.04%) students were in the most adherent group, intending to follow all guidelines, 306 (46.21%) in the moderately-adherent group, and 111 (15.75%) in the least-adherent group, rarely intending to follow any measure, with adherence negatively correlated with seropositivity of 25.4%, 32.2% and 37.7%, respectively. Moving all individuals in an SIR model into the most adherent group resulted in a 76-93% reduction in seroprevalence, dependent on assumed assortativity. The potential impact of increasing PHM adherence was limited by the substantial exposure risk in the large proportion of students already following all PHMs.

## Background

Within epidemiology, the importance of heterogeneity, whether that host, population, statistical, or environmental, has long been recognized [1–5]. For example, when designing targeted interventions, it is crucial to understand and account for differences that may exist within populations [6–8]. These differences can present in a variety of forms: heterogeneity in susceptibility, transmission, response to guidance, and treatment effects etc.; all of which affect the dynamics of an infectious disease [1,2,6,9–14]. While heterogeneity may exist on a continuous spectrum, it can be difficult to incorporate into analysis and interpretation, so individuals are often placed in discrete groups according to a characteristic that aims to represent the true differences [15–19]. When examining optimal influenza vaccination policy in the United Kingdom, Baguelin et al. [20] classified individuals within one of seven age groups. Explicitly accounting for, and grouping, individuals by whether they inject drugs can help target interventions to reduce human immunodeficiency virus (HIV) and Hepatitis C Virus incidence [21]. Similarly, epidemiological models have demonstrated the potential for HIV pre-exposure prophylaxis to reduce racial disparities in HIV incidence [22]. Therefore, heterogeneity can be used to inform more complete theories of change, increasing intervention effectiveness [23]

When discretizing a population for the purposes of inclusion within a mechanistic model, three properties need to be defined: 1) the number of groups, 2) the size of the groups, and 3) the differences between the groups. Typically, as seen in the examples above, demographic data is used e.g., age, sex, race, ethnicity, socio-economic status, etc., often in conjunction with the contact patterns and rates [7,9,15,17,20,22,24]. There are several reasons for this: the data is widely available, and therefore can be applied almost universally; it is easily understandable; and there are clear demarcations of the groups, addressing properties 1) and 2). However, epidemiological models often aim to assess the effects of heterogeneity with respect to infection, e.g., “how does an individual’s risk tolerance affect their risk of infection for influenza?”. When addressing questions such as these, demographic data does not necessarily provide a direct link between the discretization method and the heterogeneous nature of the exposure and outcome, particularly if behavioral mechanisms are a potential driver. Instead, it relies on assumptions and proxy measures e.g., an individual’s age approximates their contact rates, which in turn approximates their risk of transmission. This paper demonstrates an alternative approach to discretizing populations for use within mechanistic models, highlighting the benefits of an interdisciplinary approach to characterize heterogeneity in a manner more closely related to the risk of infection.

In early 2020, shortly after the World Health Organization (WHO) declared the SARS-CoV-2 outbreak a public health emergency of international concern [25], universities across the United States began to close their campuses and accommodations, shifting to remote instruction [26,27]. By Fall 2020, academic institutions transitioned to a hybrid working environment (in-person and online), requiring students to return to campuses [28–30]. In a prior paper [31] we documented the results of a large prospective serosurvey conducted in State College, home to The Pennsylvania State University (PSU) University Park (UP) campus. We examined the effect of 35,000 returning students (representing a nearly 20% increase in the county population [32]) on the community infection rates, testing serum for the presence of anti-Spike Receptor Binding Domain (S/RBD) IgG, indicating prior exposure [33]. Despite widespread concern that campus re-openings would lead to substantial increases in surrounding community infections [28,34,35], very little sustained transmission was observed between the two geographically coincident populations [31].

Given the high infection rate observed among the student body (30.4% seroprevalence), coupled with the substantial heterogeneity in infection rates between the two populations, we hypothesized that there may be further variation in exposure within the student body, resulting from behavioral heterogeneity. Despite extensive messaging campaigns conducted by the University [36], it is unlikely that all students equally adhered to public health guidance regarding SARS-CoV-2 transmission prevention. We use students’ responses to the behavioral survey to determine and classify individuals based on their intention to adhere to public health measures (PHMs). We then show that these latent classes are correlated with SARS-CoV-2 seroprevalence. Finally, we parameterize a mechanistic model of disease transmission within and between these groups, and explore the impact of public health guidance campaigns, such as those conducted at PSU [36]. We show that interventions designed to increase student compliance with PHMs would likely reduce overall transmission, but the relatively high initial compliance limits the scope for improvement via PHM adherence alone.

## Methods

### Design, Setting, and Participants

This research was conducted with PSU Institutional Review Board approval and in accordance with the Declaration of Helsinki, and informed consent was obtained for all participants. The student population has been described in detail previously [31], but in brief, students were eligible for the student cohort if they were: ≥ 18 years old; fluent in English; capable of providing their own consent; residing in Centre County at the time of recruitment (October 2020) with the intention to stay through April 2021; and officially enrolled as PSU UP students for the Fall 2020 term. Upon enrollment, students completed a behavioral survey in REDCap [37] to assess adherence and attitudes towards public health guidance, such as attendance at gatherings, travel patterns, and non-pharmaceutical interventions. Shortly after, they were scheduled for a clinic visit where blood samples were collected. Students were recruited via word-of-mouth and cold-emails.

### Outcomes

The primary outcome was the presence of S/RBD IgG antibodies, measured using an indirect isotype-specific (IgG) screening ELISA developed at PSU [38]. An optical density (absorbance at 450 nm) higher than six standard deviations above the mean of 100 pre-SARS-CoV-2 samples collected in November 2019, determined a threshold value of 0.169 for a positive result. Comparison against virus neutralization assays and RT-PCR returned sensitivities of 98% and 90%, and specificities of 96% and 100%, respectively [39]. Further details in the Supplement of the previous paper [31].

### Statistical Methods

To identify behavioral risk classes, we fit a range of latent class analysis (LCA) models (two to seven class models) to the student’s behavioral survey responses, using the poLCA package [40] in the R programming language, version 4.3.3 (2024-02-29) [41]. We considered their answers regarding the frequency with which they intended to engage in the following behaviors to be *a priori* indicators of behavioral risk tolerance: wash hands with soap and water for at least 20s; wear a mask in public; avoid touching their face with unwashed hands; cover cough and sneeze; stay home when ill; seek medical attention when experiencing symptoms and call in advance; stay at least 6 feet (about 2 arms lengths) from other people when outside of their home; and, stay out of crowded places and avoid mass gatherings of more than 25 people. The behavioral survey collected responses on the Likert scale of: Never, Rarely, Sometimes, Most of the time, and Always. For all PHMs, Always and Most of the time accounted for > 80% of responses (with the exception of intention to stay out of crowded places and avoid mass gatherings, where Always and Most of the time accounted for 78.8% of responses). To reduce the parameter space of the LCA and minimize overfitting, the behavioral responses were recoded as Always and Not Always. Measures of SARS-CoV-2 exposure e.g., IgG status, were not included in the LCA model fitting, as they reflect the outcome of interest. We focused on responses regarding intention to follow behaviors because this information can be feasibly collected during a public health campaign for a novel or emerging outbreak; it has also been shown that intentions are well-correlated with actual behaviors for coronavirus disease 2019 (COVID-19) public health guidelines, as well as actions that have short-term benefits [42,43]. We examined the latent class models using Bayesian Information Criterion, which is a commonly recommended as part of LCA model evaluation [44,45], to select the model that represented the best balance between parsimony and maximal likelihood fit.

Using the best-fit LCA model, we performed multivariate logistic regression of modal class assignment against IgG seropositivity to assess the association between the latent classes and infection. This “three-step” approach is recommended over the “one-step” LCA model fit that includes the outcome of interest as a covariate in the LCA model [45,46]. The following variables were determined a priori to be potential risk factors for exposure [31]: close proximity (6 feet or less) to an individual who tested positive for SARS-CoV-2; close proximity to an individual showing key COVID-19 symptoms (fever, cough, shortness of breath); lives in University housing; ate in a restaurant in the past 7 days; ate in a dining hall in the past 7 days; only ate in their room/apartment in the past 7 days; travelled in the 3 months prior to returning to campus; and travelled since returning to campus for the Fall term. Variables relating to attending gatherings were not included in the logistic regression due to overlap with intention variables of the initial LCA fit. Missing variables were deemed “Missing At Random” and imputed using the mice package [47], as described in the supplement of the previous paper [31].

We parameterized a deterministic compartmental Susceptible-Infected-Recovered (SIR) model using approximate Bayesian computation (ABC) against the seroprevalence within each latent class. The recovery rate was set to 8 days. Diagonal values of the transmission matrix were constrained such that *β*_*HH*_ ≤ *β*_*MM*_ ≤ *β*_*LL*_ (*H* represents high-adherence to public health guidelines, and *M* and *L* represent medium- and low-adherence, respectively), with the following parameters fit: the transmission matrix diagonals, a scaling factor for the off-diagonal values (*ϕ*), and a scaling factor for the whole transmission matrix (*ρ*). The off-diagonal values are equal to a within-group value (diagonal) multiplied by a scaling factor (*ϕ*). This scaling factor can either multiply the within-group beta value of the source group (e.g., *β*_*HL*_ = *ϕ* ⋅ *β*_*LL*_; Eq. 1A), or the recipient group (e.g., *β*_*LH*_ = *ϕ* ⋅ *β*_*LL*_; Eq. 1B), each with a different interpretation.

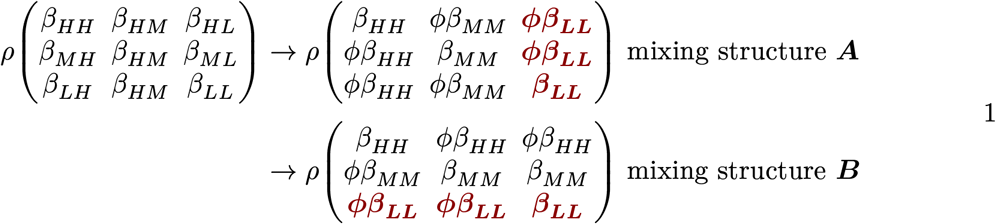

The former assumes that between-group transmission is dominated by the transmissibility of the source individuals, implying that adherence to the PHMs primarily prevents onwards transmission, rather than protecting against infection. The latter assumes that between-group transmission is dominated by the susceptibility of the recipient individuals, implying that adherence to the PHMs primarily prevents infection, rather than protecting against onwards transmission. A range of between-group scaling values (*ϕ*) were simulated to perform sensitivity analysis for the degree of assortativity. Results are only shown for matrix structure ***A***, but alternative assumptions about between-group mixing can be found in the supplement (Supplemental Figures 1-4). To examine the effect of an intervention to increase PHM adherence, we redistributed a proportion of low- and medium adherence individuals to the high adherence latent class, i.e., a fully effective intervention is equivalent to a single-group SIR model of high adherent individuals. Model fitting and simulation was conducted using the Julia programming language, version 1.10.5 [48].

## Results

### Demographics

Full details can be found in the prior paper [31], but briefly: 1410 returning students were recruited, 725 were enrolled, and 684 students completed clinic visits for serum collection between 26 October and 21 December 2020. Of these, 673 students also completed the behavioral survey between 23 October and 8 December 2020. The median age of the participants was 20 years (IQR: 19-21), 64.5% identified as female and 34.6% as male, and 81.9% identified as white. A large proportion (30.4%) were positive for IgG antibodies, and 93.5% (100) of the 107 students with a prior positive test reported testing positive only after their return to campus.

### LCA Fitting

Of the 673 participants, most students intended to always mask (81.0%), always cover their coughs/ sneezes (81.9%), and always stay home when ill (78.2%) (Table 1). Two of the least common intentions were social distancing by maintaining a distance of at least 6 feet from others outside of their home, avoiding crowded places and mass gatherings > 25 people (43.4% and 53.1% respectively), and avoiding face-touching with unwashed hands (43.5%).

**Table 1:** Participants’ intention to always or not always follow 8 public health measures.

| **Intention to always:** | **Always** | **Not Always** |
| --- | --- | --- |
| Avoid face-touching with unwashed hands | 293 (43.54%) | 380 (56.46%) |
| Cover cough and sneeze | 551 (81.87%) | 122 (18.13%) |
| Seek medical attention when have symptoms and call in advance | 480 (71.32%) | 193 (28.68%) |
| Stay at least 6 feet (about 2 arms lengths) from other people when outside of home. | 292 (43.39%) | 381 (56.61%) |
| Stay home when ill | 526 (78.16%) | 147 (21.84%) |
| Stay out of crowded places and avoid mass gatherings > 25 people | 357 (53.05%) | 316 (46.95%) |
| Tested for COVID-19 twice or more | 544 (80.83%) | 129 (19.17%) |
| Wash hands often with soap and water for at least 20 seconds. | 434 (64.49%) | 239 (35.51%) |
| Wear a face cover (mask) in public | 545 (80.98%) | 128 (19.02%) |

The four- and the three-class LCA models had the lowest BIC respectively (Table 2). Examining the four-class model, there was minimal difference in the classification of individuals, relative to the three-class model. In the four-class model, the middle class (of the three-class model) was split into two groups with qualitatively similar class-conditional item response probabilities i.e., conditional on class membership, the probability of responding “Always” to a given question, except for hand washing and avoiding face-touching with unwashed hands (Supplemental Tables 1 & 2).

**Table 2:** Log likelihood, AIC, and BIC of two to seven class LCA model fits.

| **Classes** | **Log Likelihood** | **Akaike Information Criterion** | **Bayesian Information Criterion** |
| --- | --- | --- | --- |
| 2 | -2895.40 | 5828.81 | 5914.53 |
| 3 | -2715.67 | 5489.35 | 5620.19 |
| 4 | -2673.50 | 5425.00 | 5600.96 |
| 5 | -2658.46 | 5414.93 | 5636.00 |
| 6 | -2647.01 | 5412.03 | 5678.22 |
| 7 | -2636.05 | 5410.10 | 5721.41 |

We fit a logistic regression model to predict binary IgG serostatus that included inferred class membership, in addition to other predictor variables we previously identified in [31]. The mean and median BIC and AIC indicated similar predictive ability of the three- and four-class LCA models (Table 3). Given these factors, the three-class model was selected for use in simulation for parsimony, requiring fewer assumptions and parameters to fit.

**Table 3:** Mean and median AIC and BIC of multiply-imputed logistic regressions for two to seven class LCA models against IgG serostatus.

| Classes | AIC (Mean) | BIC (Mean) | AIC (Median) | BIC (Median) |
| --- | --- | --- | --- | --- |
| 2 | 794.33 | 839.44 | 794.18 | 839.29 |
| 3 | 794.29 | 843.92 | 794.23 | 843.86 |
| 4 | 797.52 | 851.66 | 797.50 | 851.64 |
| 5 | 799.69 | 858.34 | 799.70 | 858.35 |
| 6 | 796.91 | 860.08 | 796.84 | 860.00 |
| 7 | 794.68 | 862.36 | 794.67 | 862.35 |

In the three-class model, approximately 15.75% of individuals were members of the group that rarely intended to always follow the PHMs, 38.04% intended to always follow all guidelines, and the remaining 46.21% mostly intended to mask, test, and manage symptoms, but not distance or avoid crowds (Table 4). We have labelled the three classes as “Low-”, “High-” and “Medium-Adherence” groups, respectively, for ease of interpretation. Examining the class-conditional item response probabilities, the Medium Adherence class had a probability of 0.88 of always wearing a mask in public, but a probability of only 0.19 of social distancing when outside of their homes, for example. Calculating the class-specific seroprevalence, the Low Adherence group had the highest infection rates (37.7%, 95% Binomial CI: 28.5-47.7%), the medium adherence the next highest (32.2%, 95% Binomial CI: 27.0-37.7%), and the most adherent group experienced the lowest infection rates (25.4%, 95% Binomial CI: 20.2-31.1%). Incorporating latent class membership into the imputed GLM model described in our previous paper (30) retained the relationship between adherence and infection. Relative to the least adherent group, the Medium Adherence group experienced a non-significant reduction in infection risk (aOR, 95% CI: 0.73, 0.45-1.18), and the most adherent group a significant reduction (aOR, 95% CI: 0.59, 0.36-0.98) (Table 5).

**Table 4:** Class-conditional item response probabilities shown in the main body of the table for a three-class LCA model, with footers indicating the size of the respective classes, and the class-specific seroprevalence.

| **Measure**<br>**Intention to Always:** | **Low Adherence** | **Medium Adherence** | **High Adherence** |
| --- | --- | --- | --- |
| Wash my hands often with soap and water for at least 20 seconds. | 0.04 | 0.57 | 0.96 |
| Wear a face cover (mask) in public | 0.13 | 0.88 | 0.99 |
| Avoid face-touching with unwashed hands | 0.00 | 0.21 | 0.86 |
| Cover cough and sneeze | 0.22 | 0.86 | 1.00 |
| Stay home when ill | 0.07 | 0.83 | 1.00 |
| Seek medical attention when have symptoms and call in advance | 0.03 | 0.70 | 0.98 |
| Stay at least 6 feet (about 2 arms lengths) from other people when outside of my home. | 0.00 | 0.19 | 0.87 |
| Stay out of crowded places and avoid mass gatherings > 25 people | 0.03 | 0.39 | 0.88 |
| Tested for COVID-19 twice or more | 0.76 | 0.82 | 0.81 |
| **Group Size** | **15.75%** | **46.21%** | **38.04%** |
| **Seroprevalence** | **37.7%** | **32.2%** | **25.4%** |

**Table 5:** Adjusted odds ratio (aOR) for risk factors of infection among the returning PSU UP student cohort.

| **Covariate (response) / reference levels** | **aOR (multiple imputation)** |
| --- | --- |
| Close proximity to known COVID-19 positive individual (yes) / no | 3.41 (2.29-5.08, p<0.001) |
| Close proximity to individual showing COVID-19 symptoms (yes) / no | 0.86 (0.58-1.29, p=0.474) |
| Lives in University housing (yes) / no | 0.90 (0.55-1.47, p=0.685) |
| Latent Class (medium adherence) / low adherence | 0.73 (0.45-1.18, p=0.203) |
| Latent Class (high adherence) / low adherence | 0.59 (0.36-0.98, p=0.043) |
| Travelled in the 3 months prior to campus arrival (yes) / no | 1.12 (0.76-1.63, p=0.57) |
| Travelled since campus arrival (yes) / no | 0.87 (0.6-1.25, p=0.447) |
| Ate in a dining hall in the past 7 days (yes) / no | 1.32 (0.76-2.29, p=0.332) |
| Ate in a restaurant in the past 7 days (yes) / no | 1.14 (0.8-1.64, p=0.465) |
| Only ate in their room in the past 7 days (yes) / no | 0.87 (0.59-1.29, p=0.499) |

### Compartmental Model

The ABC distance distributions indicated that near-homogeneous levels of between-group mixing better fit the data (Figure 1). After model parameterization, we examined the effect of increasing adherence to public health guidance. Moving all individuals into the High Adherence class resulted in a 76-93% reduction in final size; when moderate between-group mixing is simulated, a fully effective intervention results in approximately 80% reduction in final seroprevalence, and when between-group mixing is as likely as within-group mixing, a 93% reduction is observed (Figure 2).

**Figure 1:**
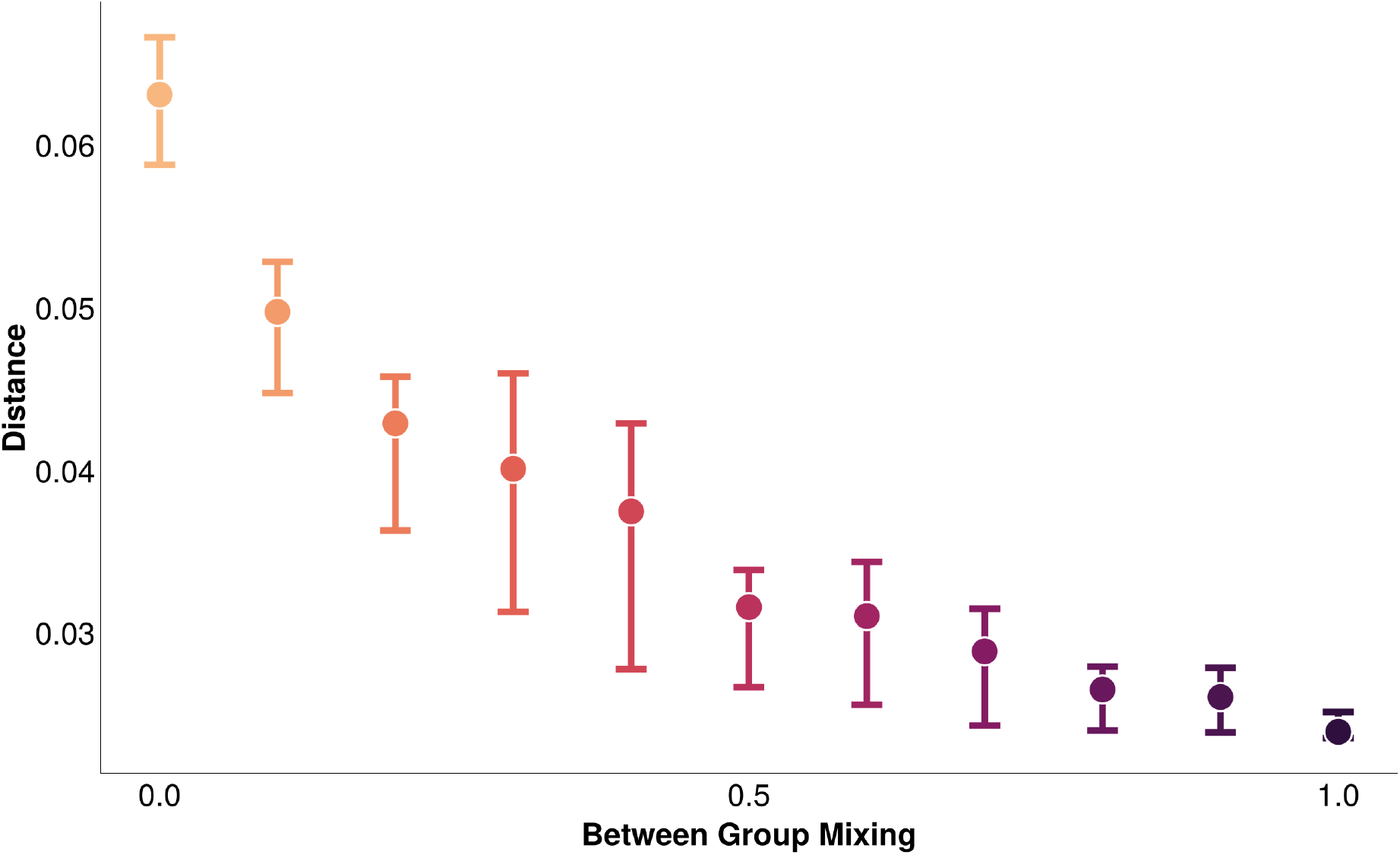
Distribution of the distance from the ABC fits, with the minimum and maximum distances illustrated by the whiskers, and the median distance by the point. Between-group mixing of 1.0 equates to between-group mixing as likely as within-group mixing

**Figure 2:**
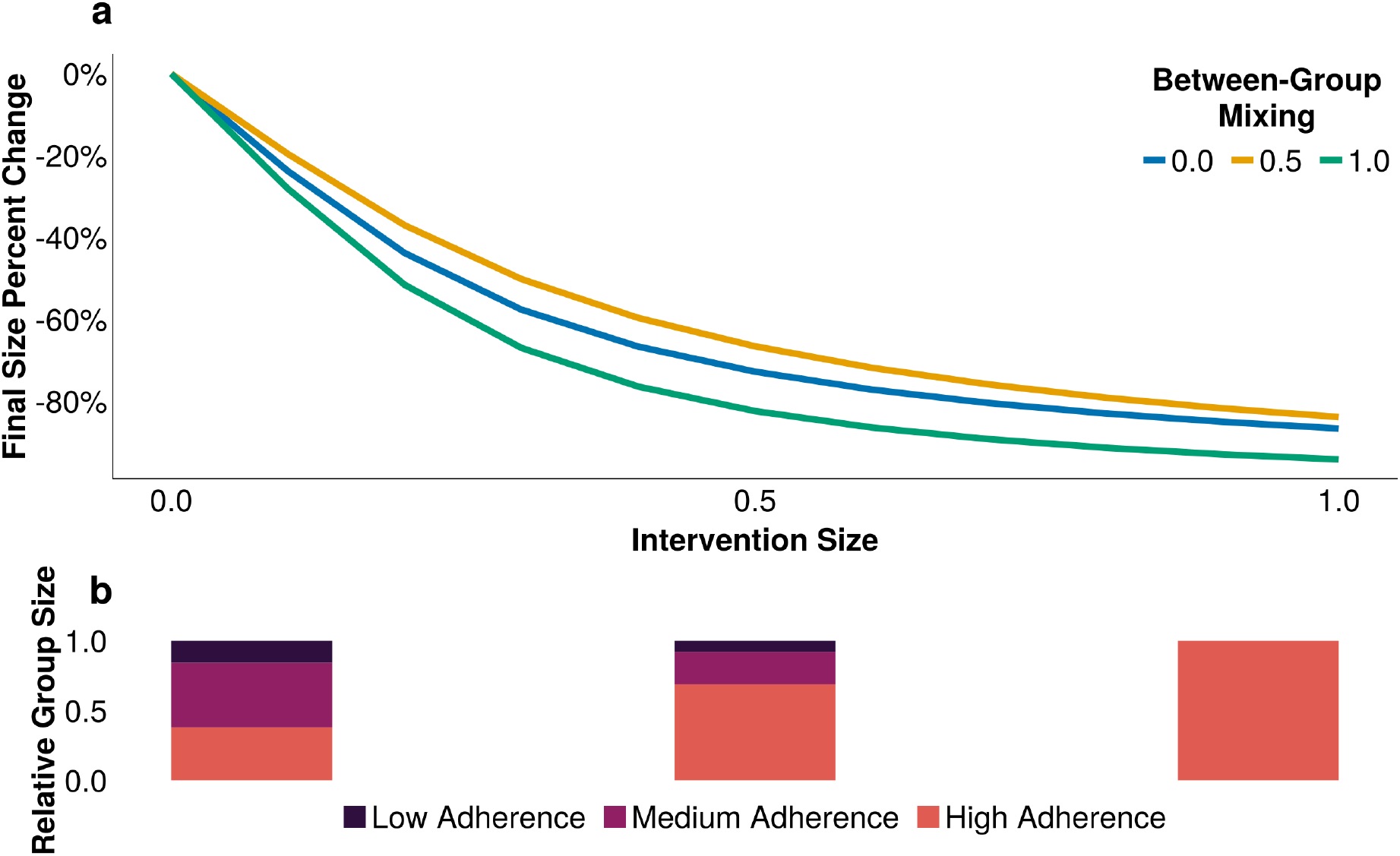
A) The reduction in final infection size across a range of intervention effectiveness (1.0 is a fully effective intervention), accounting for a range of assortativity. Between-group mixing of 1.0 equates to between-group mixing as likely as within-group mixing; B) The relative distribution of group sizes at three levels of intervention effectiveness (0.0, 0.5, 1.0)

## Discussion

In this interdisciplinary analysis, we collected behavioral data from surveys and integrated it with serosurveillance results. This approach allowed us to use LCA to categorize a population’s transmission potential with measures related to risk tolerance and behavior. The LCA model was fit without inclusion of infection status data, but class membership was correlated with IgG seroprevalence. The classes that were the most adherent to PHMs experienced the lowest infection rates, and the least adherent exhibited the highest seroprevalence.

Although a four-class LCA model was a marginally better fit for the data, there were not substantial differences in class assignment relative to the three-class LCA model. The three-class model was selected for use in simulation for parsimony, requiring fewer assumptions and parameters to fit. Upon parametrizing the compartmental model, smaller ABC distance values were observed for moderate to high levels of between-group mixing, implying some degree of assortativity in our population, though the exact nature cannot be determined from our data. Examining the three classes, 38% of individuals already intended to always follow all PHMs. As a result, only 62% of the study population could have their risk reduced with respect to the PHMs surveyed. Further, the infection rates observed in the High Adherence group indicates that even a perfectly effective intervention aimed at increasing adherence to non-pharmaceutical PHMs (i.e., after the intervention, all individuals always followed every measure) would not eliminate transmission in a population, an observation that aligns with prior COVID-19 research [49–52]. The extent to which the infection in the High Adherence group is a result of mixing with lower adherence classes cannot be explicitly described, but the sensitivity analysis allows for an exploration of the effect and ABC fits suggest near-homogeneous mixing occurred. Varying the structure of the transmission matrix yielded very similar quantitative and qualitative results (Supplemental Figures 1-4).

Examining the impact of increasing adherence to PHMs (modeled as increasing the proportion of the population in the High Adherence class), a fully effective intervention saw between a 76-93% reduction in the final size of the simulation outbreak. The small but appreciable dependence of the reduction’s magnitude on the degree of between-group mixing can be explained as such: with higher levels of between-group mixing, the initial SIR parameterization results in lower transmission parameters for the High-High adherence interactions, as more infections in the High Adherence group originate from interactions with Low and Medium Adherence individuals. Increasing adherence, therefore, results in a greater reduction of the overall transmission rate than in simulations with less assortativity.

### Limitations and Strengths

The student population was recruited using convenience sampling, and therefore may not be representative of the wider population. Those participating may have been more cognizant and willing to follow public health guidelines. Similarly, because of the University’s extensive messaging campaigns and efforts to increase access to non-pharmaceutical measures [36], such as lateral flow and polymerase-chain reaction diagnostic tests, the students likely had higher adherence rates than would be observed in other populations. However, these limitations are not inherent to the modeling approach laid out, and efforts to minimize them would likely result in stronger associations and conclusions due to larger differences in the latent behavioral classes and resulting group infection rates.

It is well known that classification methods, like LCA, can lead to the “naming fallacy” [44], whereby groups are assigned and then specific causal meaning is given to each cluster, affecting subsequent analyses and interpretation of results. In this paper, this effect is reduced by virtue of the analysis plan being pre-determined, and the relationship with the outcome showing a positive association with the classes in the mechanistically plausible direction (i.e., increasing adherence to PHMs results in reduced infection rates). Our decision to conduct the simulation analysis with the three-class model was, in part, to avoid the potential bias that would arise from naming or assigning an order to the two intermediate risk groups.

Despite these limitations, this work presents a novel application of a multidisciplinary technique, outlining how alternate data sources can guide future model parameterization and be incorporated into traditional epidemiological analysis, particularly within demographically homogeneous populations where there is expected or observed heterogeneity in transmission dynamics. This is particularly important in the design of interventions that aim to target individual behaviors, allowing the categorization of populations into dynamically-relevant risk groups and aiding in the efficient use of resources through targeted actions. Future research should consider including perceived agency and efficacy for PHM adherence.

## Supporting information

Supplemental Results

## Data Availability

The datasets generated during and/or analyzed during the current study are not publicly available as they contain personally identifiable information, but are available from the corresponding author on reasonable request.

## Additional Information

### Data Access, Responsibility, and Analysis

Callum Arnold and Dr. Matthew J. Ferrari had full access to all the data in the study and take responsibility for the integrity of the data and the accuracy of the data analysis. Callum Arnold and Dr. Matthew J. Ferrari (Department of Biology, Pennsylvania State University) conducted the data analysis.

## Author Contributions

*Conceptualization:* CA, MJF

*Data curation:* CA, MJF

*Formal analysis:* CA, MJF

*Funding acquisition:* MJF

*Investigation:* NB, CE, MS, SS, SK, VS

*Methodology:* CA, NB, MJF

*Project administration:* MJF

*Software:* CA, MJF

*Supervision:* MJF

*Validation:* CA, MJF

*Visualization:* CA, MJF

*Writing - original draft:* CA

*Writing - review and editing:* all authors.

## Acknowledgements

1. Florian Krammer, Mount Sinai, USA for generously providing the transfection plasmid pCAGGS-RBD
2. Scott E. Lindner, Allen M. Minns, Randall Rossi produced and purified RBD
3. The D4A Research Group: Dee Bagshaw, Clinical & Translational Science Institute, Cyndi Flanagan, Clinical Research Center and the Clinical & Translational Science Institute; Thomas Gates, Social Science Research Institute; Margeaux Gray, Dept. of Biobehavioral Health; Stephanie Lanza, Dept. of Biobehavioral Health and Prevention Research Center; James Marden, Dept. of Biology and Huck Institutes of the Life Sciences; Susan McHale, Dept. of Human Development and Family Studies and the Social Science Research Institute; Glenda Palmer, Social Science Research Institute; Connie J. Rogers, Dept. of Nutritional Sciences; Rachel Smith, Dept. of Communication Arts and Sciences and Huck Institutes of the Life Sciences; and Charima Young, Penn State Office of Government and Community Relations.
4. The authors thank the following for their assistance in the lab: Sophie Rodriguez, Natalie Rydzak, Liz D. Cambron, Elizabeth M. Schwartz, Devin F. Morrison, Julia Fecko, Brian Dawson, Sean Gullette, Sara Neering, Mark Signs, Nigel Deighton, Janhayi Damani, Mario Novelo, Diego Hernandez, Ester Oh, Chauncy Hinshaw, B. Joanne Power, James McGee, Riëtte van Biljon, Andrew Stephenson, Alexis Pino, Nick Heller, Rose Ni, Eleanor Jenkins, Julia Yu, Mackenzie Doyle, Alana Stracuzzi, Brielle Bellow, Abriana Cain, Jaime Farrell, Megan Kostek, Amelia Zazzera, Sara Ann Malinchak, Alex Small, Sam DeMatte, Elizabeth Morrow, Ty Somberger, Haylea Debolt, Kyle Albert, Corey Price, Nazmiye Celik

