## Supplemental Results for "The Maximal Expected Benefit of SARS-CoV-2 Interventions Among University Students: A Simulation Study Using Latent Class Analysis"

### LCA Model Fitting

| **Measure<br>Intention to Always:** | **Low Adherence** | **Low-Medium Adherence** | **Medium-High Adherence** | **High Adherence** |
|---|---|---|---|---|
| Wash my hands often with soap and water for at least 20 seconds. | 0.04 | 0.38 | 0.93 | 0.95 |
| Wear a face cover (mask) in public | 0.11 | 0.88 | 0.88 | 0.99 |
| Avoid face-touching with unwashed hands | 0.00 | 0.00 | 0.62 | 0.85 |
| Cover cough and sneeze | 0.22 | 0.77 | 1.00 | 1.00 |
| Stay home when ill | 0.06 | 0.82 | 0.85 | 0.99 |
| Seek medical attention when have symptoms and call in advance | 0.02 | 0.68 | 0.75 | 0.98 |
| Stay at least 6 feet (about 2 arms lengths) from other people when outside of my home. | 0.00 | 0.22 | 0.10 | 0.92 |
| Stay out of crowded places and avoid mass gatherings > 25 people | 0.02 | 0.46 | 0.23 | 0.92 |
| Tested for COVID-19 twice or more | 0.76 | 0.81 | 0.84 | 0.81 |
| **Group Size** | 13.82% | 30.91% | 16.49% | 38.78% |
| **Seroprevalence** | 35.50% | 31.20% | 36.00% | 25.70% |

*Supplemental Table 1: Class-conditional item response probabilities shown in the main body of the table for a four-class LCA model, with footers indicating the size of the respective classes, and the class-specific seroprevalence*

### Matrix Structure Sensitivity Analysis

In the main body of the text, we present the results for the three-class model that corresponds to a scenario where public health measures (PHMs) reduce onwards risk of transmission (Supplemental Eq 1A), rather than conferring protection for the practitioner (Supplemental Eq 1B). Another alternative uses a single scaled value of $\beta_{LL}$, representing all between-group interactions experiencing the same risk of transmission that is a fraction of the transmission observed between Low Adherence individuals (Supplemental Eq 1C).

$$
\begin{aligned}
\rho \begin{pmatrix} \beta_{HH} & \beta_{HM} & \beta_{HL} \\ \beta_{MH} & \beta_{HM} & \beta_{ML} \\ \beta_{LH} & \beta_{HM} & \beta_{LL} \end{pmatrix} &\to \rho \begin{pmatrix} \beta_{HH} & \phi\beta_{MM} & \phi\beta_{LL} \\ \phi\beta_{HH} & \beta_{MM} & \phi\beta_{LL} \\ \phi\beta_{HH} & \phi\beta_{MM} & \beta_{LL} \end{pmatrix} \text{ mixing structure } \boldsymbol{A} \\
&\to \rho \begin{pmatrix} \beta_{HH} & \phi\beta_{HH} & \phi\beta_{HH} \\ \phi\beta_{MM} & \beta_{MM} & \beta_{MM} \\ \phi\beta_{LL} & \phi\beta_{LL} & \beta_{LL} \end{pmatrix} \text{ mixing structure } \boldsymbol{B} \\
&\to \rho \begin{pmatrix} \beta_{HH} & \phi\beta_{LL} & \phi\beta_{LL} \\ \phi\beta_{LL} & \beta_{MM} & \phi\beta_{LL} \\ \phi\beta_{LL} & \phi\beta_{LL} & \beta_{LL} \end{pmatrix} \text{ mixing structure } \boldsymbol{C}
\end{aligned} \tag{1}
$$

Below are results for alternative scenarios, which show qualitatively similar results to the main body of the text, albeit with a wider distribution in the Approximate Bayesian Computation distance metrics.

***Eq 1B (PHMs Confer Protection)***

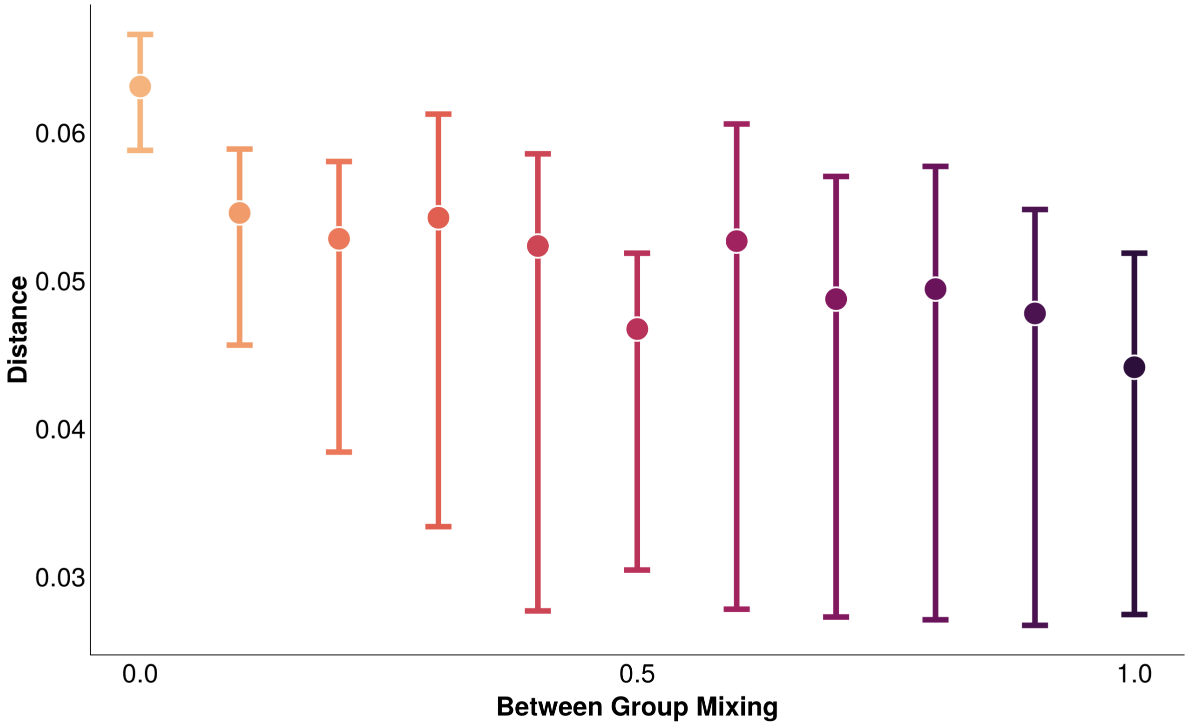


*Figure 1: PHMs confer protection to the practitioner. Distribution of the distance from the ABC fits, with the minimum and maximum distances illustrated by the whiskers, and the median distance by the point. Between-group mixing of 1.0 equates to between-group mixing as likely as within-group mixing*

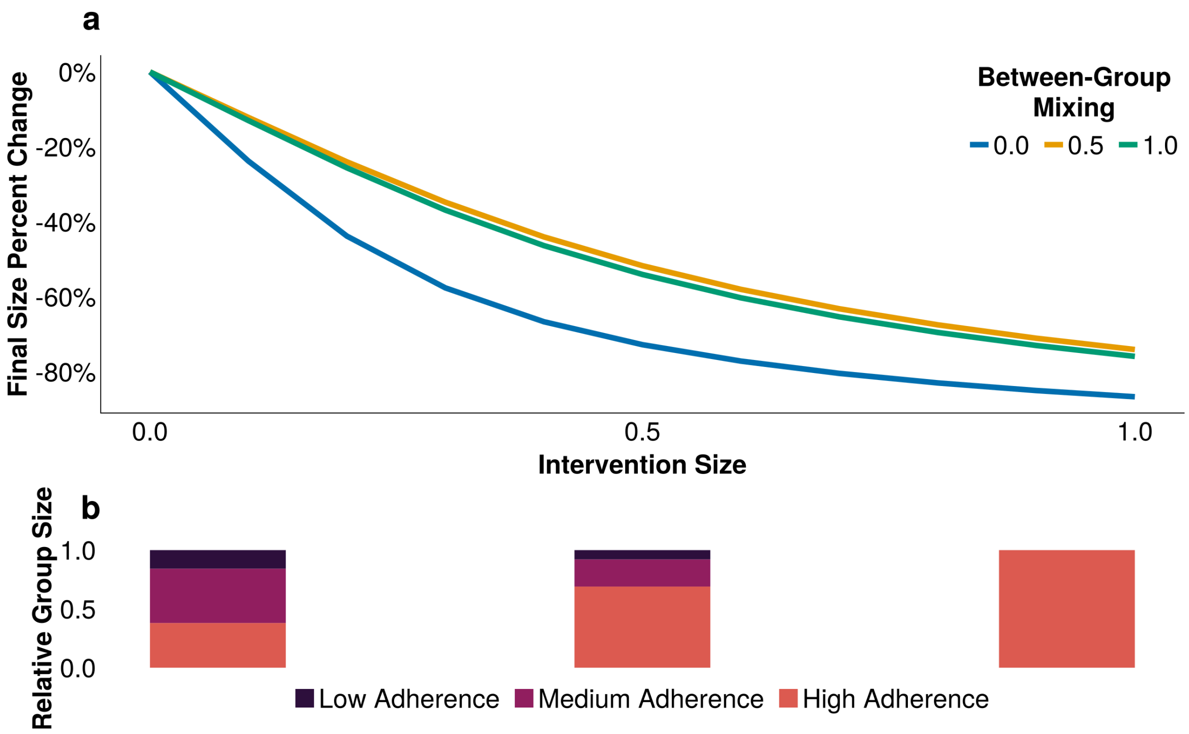


*Figure 2: PHMs confer protection to the practitioner. A) The reduction in final infection size across a range of intervention effectiveness (1.0 is a fully effective intervention), accounting for a range of assortativity. Between-group mixing of 1.0 equates to between-group mixing as likely as within-group mixing; B) The relative distribution of group sizes at three levels of intervention effectiveness (0.0, 0.5, 1.0)*

### 14 *Eq 1C (Identical Off-Diagonal Values)*

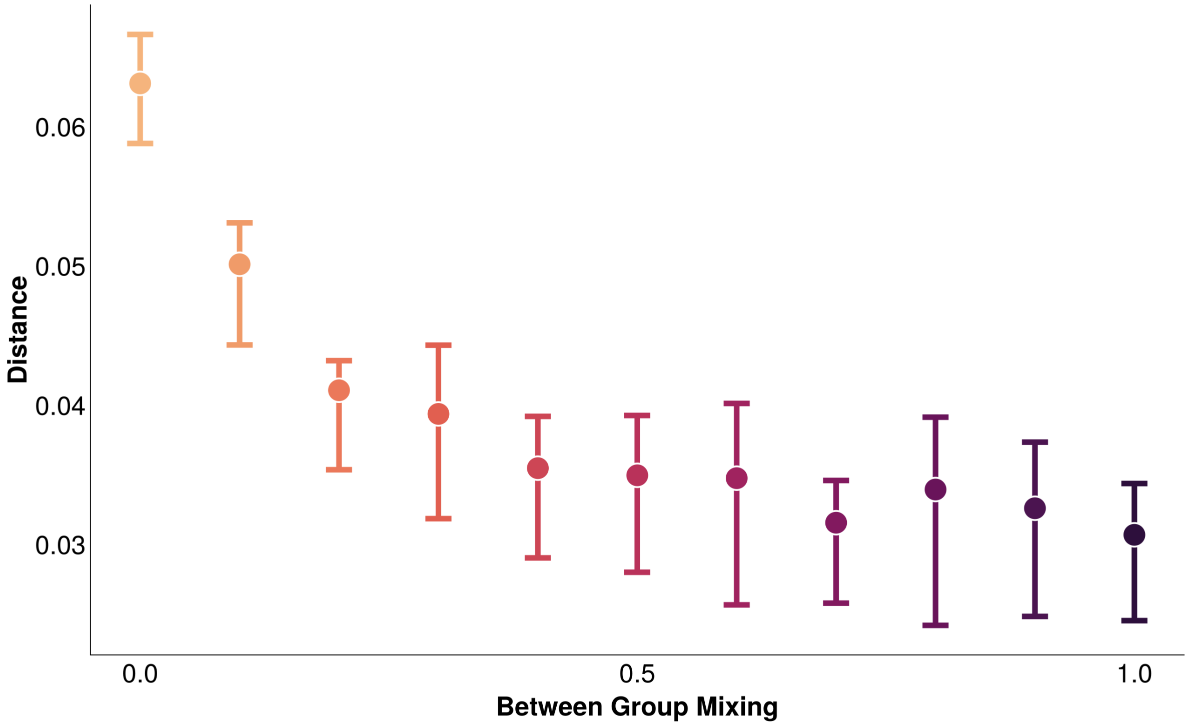


*Figure 3: Identical off-diagonal values. Distribution of the distance from the ABC fits, with the minimum and maximum distances illustrated by the whiskers, and the median distance by the point. Between-group mixing of 1.0 equates to between-group mixing as likely as within-group mixing*

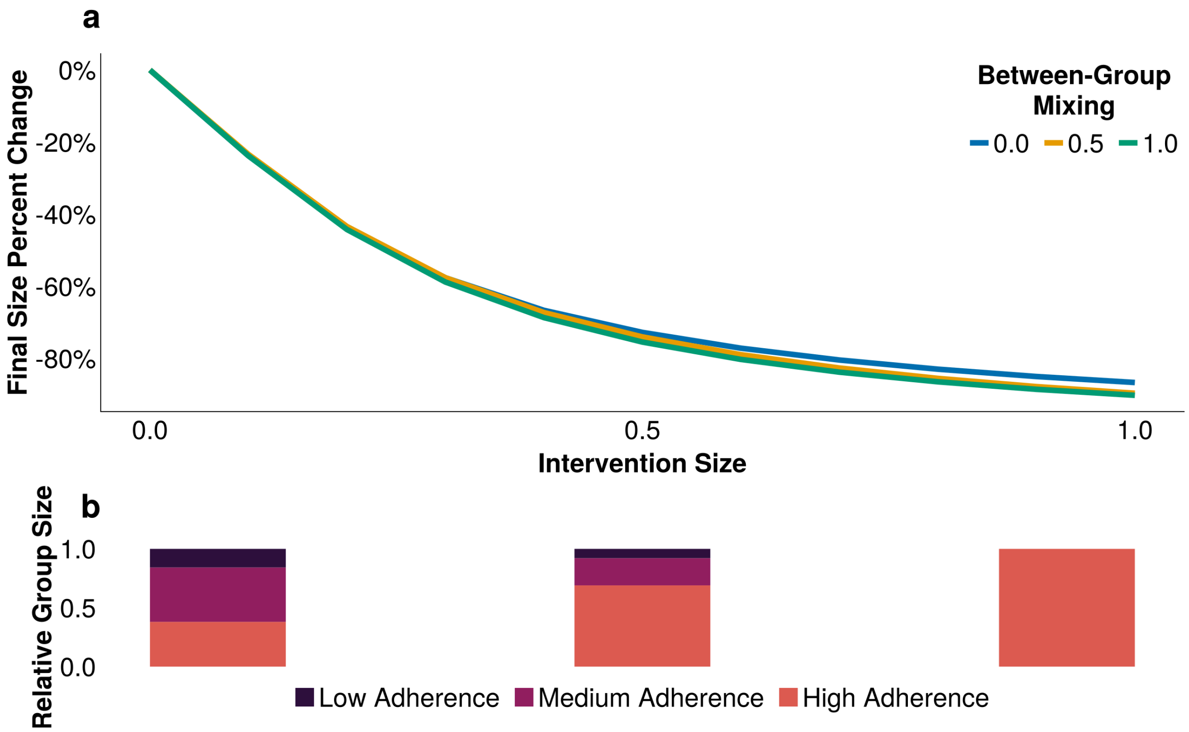


*Figure 4: Identical off-diagonal values. A) The reduction in final infection size across a range of intervention effectiveness (1.0 is a fully effective intervention), accounting for a range of assortativity. Between-group mixing of 1.0 equates to between-group mixing as likely as within-group mixing; B) The relative distribution of group sizes at three levels of intervention effectiveness (0.0, 0.5, 1.0)*
